# Transcribing multilingual radiologist-patient dialogue into mammography reports using AI: A step towards patient-centric radiology

**DOI:** 10.1101/2025.03.26.25324592

**Authors:** Amit Gupta, Ashish Rastogi, Neha Rani, Mohak Narang, Krithika Rangarajan

## Abstract

**Background:** Radiology reports are primarily designed for healthcare professionals, often containing complex medical terminology hindering patients from understanding their diagnostic results. This communication gap is especially pronounced in non-English-speaking regions. AI-driven transcription and report generation, leveraging automated speech recognition (ASR) and large language models (LLMs), could enable patient-centered, accessible reporting from radiologist-patient conversations in vernacular language.

**Purpose:** To evaluate the feasibility of AI-driven transcription and automated mammography report generation from simulated radiologist-patient conversations in vernacular language, assessing transcription accuracy, report concordance, error patterns, and time efficiency.

**Materials and Methods:** A curated dataset of 50 mammograms was retrospectively selected from the Picture Archiving and Communication System (PACS) of our department. Simulated radiologist-patient conversations, conducted in vernacular Hindi, were recorded and transcribed using the OpenAI Whisper large-v2 ASR model. Four transcriptions per conversation were generated at different temperatures (0, 0.3, 0.5, 0.7) to maximize information capture. Structured mammography reports were generated from the transcriptions using GPT-4o, guided by detailed prompt instructions. Reports were reviewed and corrected by a radiologist, and AI performance was assessed through word error rate (WER), character error rate (CER), report concordance rates, error analysis, and time efficiency metrics.

**Results:** The lowest WER (0.577) and CER (0.379) were observed at temperature 0. The overall mean concordance rate between AI-generated and radiologist-edited reports was 0.94, with structured fields achieving higher concordance than descriptive fields. Errors were present in 50% of AI-generated reports, predominantly missed and incorrect information, with a higher error rate in malignant cases. The mean time for AI-driven report generation was 207.4 seconds, with radiologist editing contributing 43.1 seconds on average.

**Conclusion:** AI-driven workflow integrating ASR and LLMs to generate structured mammography reports from radiologist-patient conversations in vernacular language, is feasible. While challenges such as privacy, validation, and scalability remain, this approach represents a significant step toward patient-centric and AI-integrated radiology practice.

## Introduction

Radiology is an essential pillar of modern healthcare, playing a crucial role in diagnosis and treatment planning. However, despite advancements in medical imaging and digital health infrastructure, radiology workflows remain largely physician-centric, with traditional reports designed primarily for healthcare professionals rather than patients (1,2). This lack of direct, structured communication between radiologists and patients is particularly problematic in non-English-speaking populations, where the technical nature of radiology reports, often written in complex medical English, creates a barrier to patient understanding (3,4).

In India, where the healthcare system follows a patient-first model, patients often receive their imaging reports directly—frequently before consulting a physician. Without immediate medical guidance, patients are left to interpret their results independently, leading to misinterpretation, unnecessary anxiety, and delays in care. Many resort to technicians, online sources, or family members to decipher findings, which may result in misinformation or diagnostic confusion. Furthermore, radiologists seldom provide patient-friendly summaries due to high caseloads and time constraints, leaving a critical gap in patient education and engagement (5,6).

Artificial intelligence (AI) offers a novel solution to this communication gap, with potential applications beyond image interpretation. Recent advancements in automated speech recognition (ASR) and large language models (LLMs) provide an opportunity to capture real-time radiologist-patient interactions and generate structured radiology reports (7–10). AI-driven transcription of vernacular-language consultations could bridge the divide between radiologists and patients, ensuring that imaging findings are presented in a manner that is both clinically accurate and easily comprehensible. By integrating ASR with language models for automated report generation, radiology workflows could become more patient-centric, reducing reliance on self-interpretation while improving accessibility for non-English-speaking populations.

This study aims to evaluate the feasibility of AI-driven transcription and automated mammography report generation from simulated radiologist-patient conversations in vernacular language. By assessing the accuracy, efficiency, and reliability of AI-generated reports, this research explores how AI can facilitate a more equitable and accessible radiology practice.

## Materials and Methods

This study was approved by the Institute Ethics Committee. A curated dataset of 50 mammograms was retrospectively selected from the Picture Archiving and Communication System (PACS) of our department. All the mammograms in our department are reported using the American College of Radiology (ACR) Breast Imaging and Reporting Data System (BI-RADS) terminology, with each scan undergoing standard double-reader assessment.

The curated dataset comprised 10 normal (BI-RADS 1), 20 benign (BI-RADS 2 or 3), and 20 malignant (BI-RADS 4 and 5) cases, ensuring the inclusion of a broad spectrum of common mammographic findings encountered in clinical practice. The dataset consisted exclusively of 2D mammographic images, with tomosynthesis images intentionally excluded. All selected mammograms were acquired using the Hologic Selenia Dimensions system.

### Simulated Radiologist-Patient Conversations

To simulate real-world patient interactions, a radiologist (with 6 years of experience in general radiology) engaged in verbal discussions regarding mammographic findings with a technical staff, who role-played as the patient. The conversations were conducted in vernacular language (Hindi), mimicking typical interactions in a clinical setting, and were audio-recorded to facilitate subsequent transcription and analysis. Structured instructions were provided to both the radiologist and the technical staff (acting as the patient) to ensure consistency and comprehensiveness in the simulated discussions.

### Instructions to the Radiologist

The radiologist was blinded to the mammographic findings prior to each conversation to mimic a real-time patient consultation scenario. A predefined list of standardized translations for the ACR BI-RADS lexicon terms into vernacular Hindi was provided to the radiologist to facilitate the use of simplified, patient-friendly language. The radiologist was instructed to comprehensively discuss all aspects of the mammogram, including:

- normal and abnormal findings, ensuring a comprehensive discussion of both reassuring and concerning aspects,
- axillary lymph node evaluation,
- interpretation of BI-RADS categories, explaining the classification in terms understandable to a layperson.

The radiologist was further instructed to engage in open-ended dialogue, allowing for a natural flow of discussion that reflected real-world patient consultations.

### Instructions to the Technical Staff (Simulated Patient)

Before each simulated consultation, the technical staff member was provided with relevant patient history, including specific breast complaints (e.g., lump, nipple discharge), annual follow-up for screening mammography, and any prior history of breast cancer or mastectomy. Additionally, the techincal staff was instructed to actively participate in the conversation by asking common patient queries that typically arise during mammography consultations. These included:

- implication of the imaging findings (e.g., whether the results were concerning),
- need for additional imaging, such as ultrasound,
- requirement for biopsy or follow-up based on the mammogram findings.

Any identifiable patient information like patient name, demographics, patient ID, etc. were not discussed in the conversations. By ensuring structured yet dynamic discussions, this simulation aimed to accurately reflect real-world radiologist-patient interactions, providing a robust dataset for evaluating AI-driven transcription and report generation.

### AI-Based Transcription and Report Generation Pipeline

The AI workflow consisted of two primary steps: ASR for speech-to-text transcription and generative AI-based report generation.

#### 1. Speech-to-text Transcription Using Whisper ASR Model

The recorded conversations were transcribed using OpenAI’s Whisper large-v2 model via an application programming interface (API). Whisper is a general-purpose speech recognition model trained on a vast and diverse audio dataset, enabling it to perform tasks such as multilingual speech recognition, translation, and language identification (11). The model includes a prompt parameter, which allows users to provide contextual text that helps guide transcription accuracy by influencing word recognition and formatting. In this study, a subset (selected due to prompt length constraints) of the predefined translated Hindi terms for BI-RADS lexicon as referred to above, were given as a prompt to the model to improve recognition of domain-specific terminology. To ensure comprehensive capture of all relevant information from the audio conversations, the transcription was executed at four different temperatures: 0 (most deterministic), 0.3, 0.5 and 0.7 (most stochastic). Thus for each case, four separate transcription versions were generated, one for each temperature setting. This approach increased the likelihood of capturing all relevant information from the audio conversation, mitigating information loss due to variations in transcription accuracy.

#### 2. Report Generation Using LLM followed by Radiologist Editing

All four transcription versions for each case were collectively processed using the Generative Pre-trained Transformer 4o (GPT-4o) accessed via API, to generate structured mammography reports (12). The AI model was provided with a detailed prompt, which included:

- Instructions to integrate information from all four versions of the Hindi audio transcripts to ensure completeness and accuracy.
- A predefined mapping of BI-RADS lexicon terms and their corresponding Hindi translations, identical to the list provided to the radiologist during the conversation.
- A standardized JavaScript Object Notation (JSON) format for the generated report as output, ensuring structured representation of key report components, including patient history, findings for the right and left breast, impression, BI-RADS category, and recommendation.

The AI-generated reports were displayed as editable JSON files in HTML format, enabling the radiologist to review and modify the content as needed before finalization. The radiologist modified the reports only to correct factual inaccuracies, ensuring the information was clinically accurate. Grammatical or language adjustments were made only if they affected the clarity or interpretation of the report’s content. The final radiologist-edited reports were also saved.

### Evaluation Metrics and Performance Analysis

The performance of the AI-based transcription and report generation system was assessed using multiple objective metrics.

#### 1. Transcription Accuracy Assessment

The recorded audio conversations were manually transcribed into Hindi text by a radiology technical staff member (with two years of experience in radiology research) to serve as a reference for evaluating AI-generated transcriptions. To quantify the accuracy of AI-generated transcriptions, the word error rate (WER) and character error rate (CER) were computed using Python scripts. WER and CER are standard metrics used in machine learning to evaluate the performance of speech recognition and transcription systems (13,14). WER measures the proportion of incorrectly transcribed words, while CER assesses errors at the character level. Lower WER and CER values indicate higher transcription accuracy. The two metrics were calculated individually for each temperature setting.

#### 2. Report Concordance Analysis

The AI-generated structured JSON outputs were compared to the final radiologist-edited reports. To calculate the concordance rate for each predefined JSON field in the reports, we determined the proportion of AI-generated fields that matched the final radiologist-approved version, excluding any fields that were intentionally left empty (e.g., in unilateral mammograms, fields corresponding to the non-imaged breast were omitted from the analysis). Concordance rates were calculated for each individual report section, as well as the overall metrics for the entire set of reports.

#### 3. Error analysis

An experienced radiologist (with 10 years of experience in general radiology) conducted a qualitative evaluation of each LLM-generated report that exhibited any discordance compared to the final radiologist-edited report. This evaluation focused on the number of errors in each discordant report, their sources, types, and severity.

##### a. Source of Error

- Transcription errors: These originate from the ASR process, where inaccuracies occur during the conversion of spoken language into text.
- LLM processing errors: These arise during the generative AI-based report generation phase, involving misinterpretations or misrepresentations by the language model.

##### b. Type of Error

- Laterality errors: Incorrect identification of the side (left vs. right) being described.
- Missed information: Omission of pertinent details present in the radiologist-patient conversation.
- Wrong information: Inclusion of incorrect details not supported by the conversation.
- False added information: Introduction of information not present in the conversation.
- Minor language errors: Errors affecting language clarity and posing minimal doubt in clinical meaning.

##### c. Severity of Error

- The errors in the AI-generated reports were categorised into three severity categories
- minor, moderate, severe - based on their potential impact on patient care.

#### 3. Time Efficiency Analysis

The time required for each of the four components for the end-to-end report generation process was recorded as follows:

- Conversation time – Duration of radiologist-technical staff interaction.
- Transcription time – Time taken by the Whisper model to generate the transcript.
- LLM processing time – Time taken for AI-based report generation.
- Editing time – Time spent by the radiologist in reviewing and modifying the AI-generated report.

### Statistical Analysis

Descriptive statistics were used to summarize WER, CER, report concordance rates, error rates and time efficiency metrics, using Python (version 3.0) scripts. The study methodology is summarised in Figure 1.

**Figure 1:**
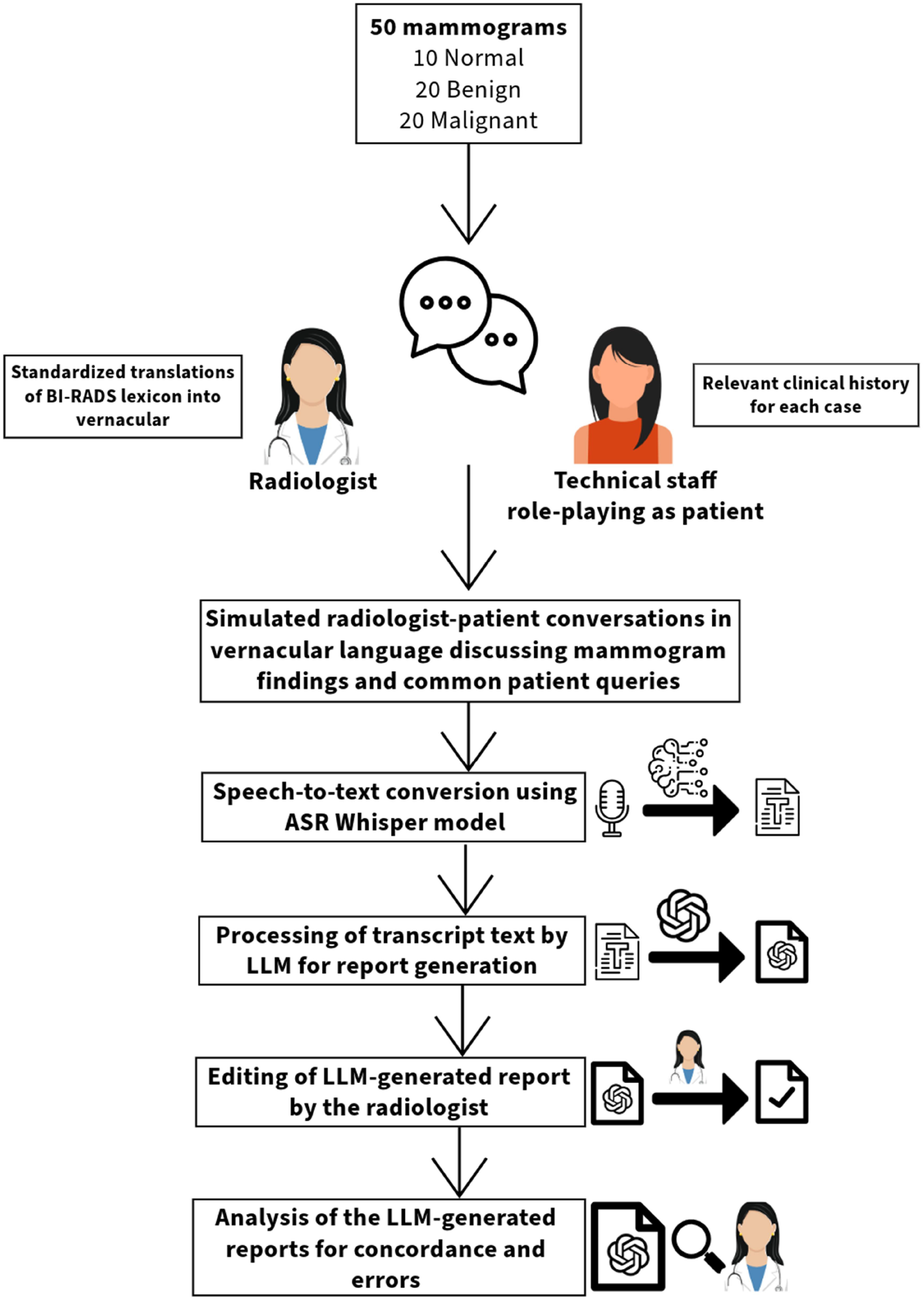
Study methodology

## Results

### 1. Transcription Accuracy

The observed WER and CER values varied across temperature settings, with the lowest values recorded at temperature 0 (WER: 0.577 ± 0.0865, CER: 0.379 ± 0.0569). Across higher temperature settings, there were slight increases in WER and CER, with values at temperature 0.3 recorded as 0.641 (± 0.0962) and 0.428 (± 0.0642), at 0.5 as 0.604 (± 0.0906) and 0.380 (± 0.0570), and at 0.7 as 0.622 (± 0.0933) and 0.395 (± 0.0593), respectively. Although lower temperature settings demonstrated marginally lower error rates, the overall differences remained minimal, suggesting that transcription accuracy remained relatively stable across different temperature settings.

### 2. Report Concordance Analysis

The overall mean concordance rate with radiologist-edited reports, across all fields in the LLM-generated JSON reports was 0.94, with values for individual fields ranging from 0.64 to 0.96. The highest concordance rates were observed for the “Recommendation” and “Comparison” fields, both achieving 0.96, indicating strong AI performance in structured elements of the report. In contrast, the “Left Breast Parenchyma” and “Right Breast Parenchyma” fields had the lowest concordance rates of 0.64 and 0.77, respectively, suggesting that descriptive, free-text components of the report were more susceptible to variability. The detailed concordance analysis for each report section and across all sections, is shown in Table 1.

**Table 1:** Concordance analysis of LLM-generated reports compared to radiologist-edited reports for each report section and across all reports.

| Section | Concordant Entries (c) | Total number of Entries (t) | Concordance Rate (c/t) |
| --- | --- | --- | --- |
| Procedure | 47 | 50 | 0.94 |
| Clinical Background | 45 | 50 | 0.90 |
| Right Breast_Breast density | 41 | 44 | 0.93 |
| Right Breast_Parenchyma | 34 | 44 | 0.77 |
| Right Breast_Areola and Subcutaneous tissues | 41 | 44 | 0.93 |
| Right Breast_Axilla | 42 | 44 | 0.95 |
| Left Breast_Breast density | 39 | 45 | 0.87 |
| Left Breast_Parenchyma | 28 | 44 | 0.64 |
| Left Breast_Areola and Subcutaneous tissues | 42 | 45 | 0.93 |
| Left Breast_Axilla | 41 | 45 | 0.91 |
| Comparison | 48 | 50 | 0.96 |
| Impression_Right Breast_Summary | 35 | 44 | 0.80 |
| Impression_Right Breast_BI-RADS Category | 40 | 44 | 0.91 |
| Impression_Left Breast_Summary | 34 | 45 | 0.76 |
| Impression_Left Breast_BI-RADS Category | 40 | 45 | 0.89 |
| Recommendation | 48 | 50 | 0.96 |
| Overall concordance rate | 645 | 683 | 0.94 |

### 3. Error Analysis

Out of the 50 cases, 25 LLM-generated reports (50%) contained one or more discrepancies as compared to radiologist-edited reports, while the remaining 25 (50%) were entirely concordant. The distribution of discordant reports varied across different case categories: 1 in 10 normal cases, 10 in 20 benign cases, and 14 in 20 malignant cases. The sources of errors were attributed to LLM processing in 12 reports, transcription in 9 reports, and both in 4 reports. A total of 36 error instances were identified and categorized by type and severity as: missed information (n=16/36), wrong information (n=13/36), laterality errors (n=3/36), false added information (n=2/36), and minor language errors (n=2/36). The majority of errors were classified as minor (n=18/36) or moderate (n=14/36), with a smaller number deemed severe (n=4/36). Detailed error analysis of the LLM-generated reports is shown in Table 2 and Table 3.

**Table 2:** Case-wise number of reports with errors and sources of these errors.

|  | **Normal (%)**<br>**N=10** | **Benign (%)**<br>**N=20** | **Malignant (%)**<br>**N=20** | **Total (%)**<br>**N=50** |
| --- | --- | --- | --- | --- |
| **Totally correct reports** | 9 (90) | 10 (50) | 6 (30) | 25 (50) |
| **Reports with errors** | 1 (10) | 10 (50) | 14 (70) | 25 (50) |
| **Source of error** |  |  |  |  |
| **LLM processing** | 0 | 9 (45) | 3 (15) | 12 (24) |
| **Transcription** | 1 (10) | 0 | 8 (40) | 9 (18) |
| **Both** | 0 | 1 (5) | 3 (15) | 4 (8) |

**Table 3:**
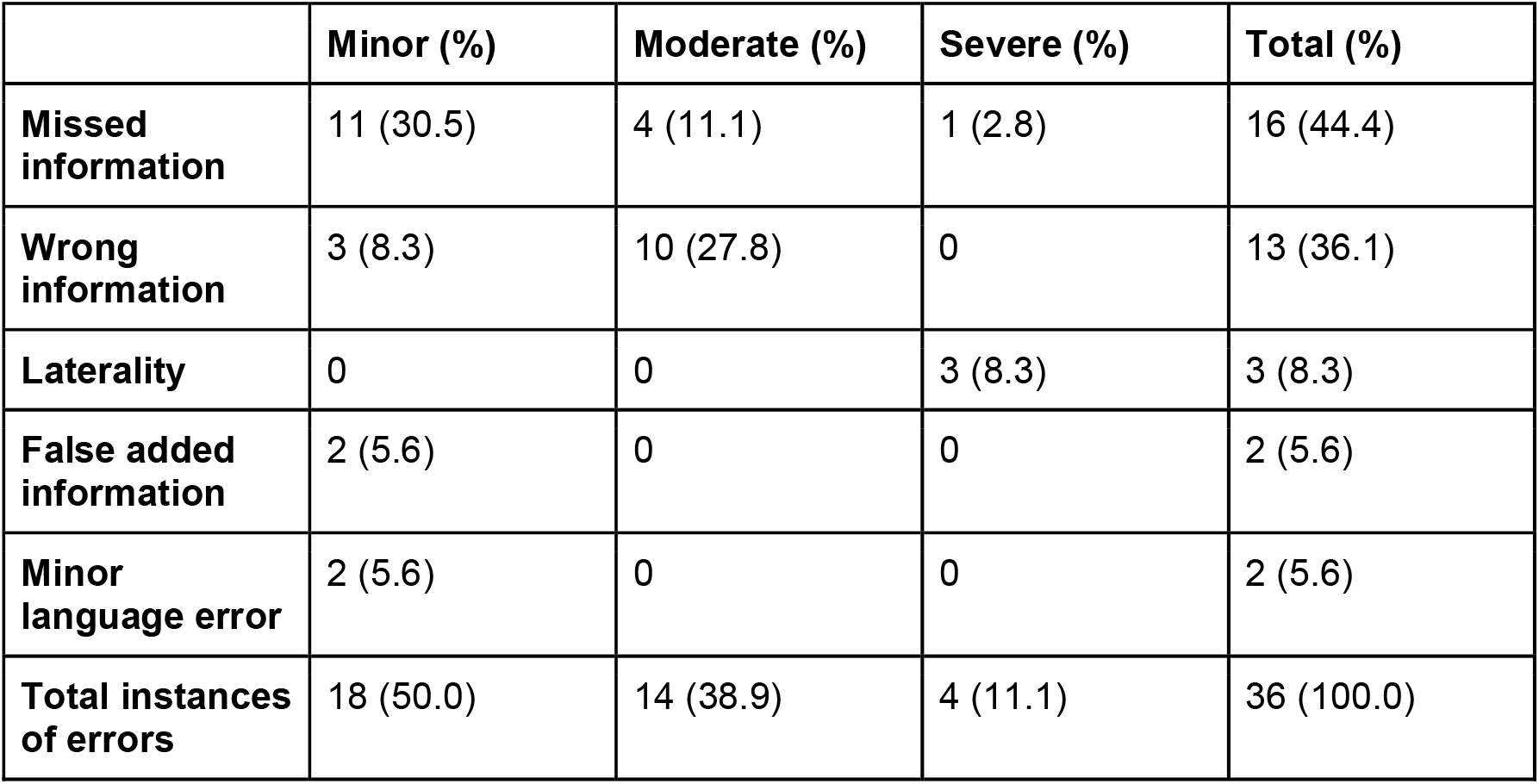
Type and severity of errors in the LLM-generated reports (N=36)

|  | **Minor (%)** | **Moderate (%)** | **Severe (%)** | **Total (%)** |
| --- | --- | --- | --- | --- |
| **Missed information** | 11 (30.5) | 4 (11.1) | 1 (2.8) | 16 (44.4) |
| **Wrong information** | 3 (8.3) | 10 (27.8) | 0 | 13 (36.1) |
| **Laterality** | 0 | 0 | 3 (8.3) | 3 (8.3) |
| **False added information** | 2 (5.6) | 0 | 0 | 2 (5.6) |
| **Minor language error** | 2 (5.6) | 0 | 0 | 2 (5.6) |
| **Total instances of errors** | 18 (50.0) | 14 (38.9) | 4 (11.1) | 36 (100.0) |

Instances of transcription-related errors included laterality mistakes, where the transcription text incorrectly identified the left breast as the right, and vice versa. Errors in documenting lesion characteristics were also noted, such as incorrect size measurements and misidentification of the quadrant location of masses. Additionally, there were cases where the transcription inaccurately documented clinical history, including omissions of patient-reported lumps or breast density, resulting in errors in the AI-generated report. LLM processing errors encompassed the omission of benign calcifications and normal breast entries in the AI-generated descriptions, despite these details being present in the transcript. There were also instances where the LLM provided incorrect breast density assessments and mass descriptors that could have been accurately interpreted from the transcript text.

Some reports contained hallucinated information, such as the addition of benign calcifications not present in the transcript. Minor language errors were also present, which, while not clinically significant, could affect the clarity of the report.

### 4. Time Efficiency Analysis

The mean total time for report generation was 207.4 (± 73.3) seconds (approximately 3.5 minutes per case), with the radiologist-patient conversation being the most time-consuming step, averaging 113.3 (± 40.9) seconds per case. The Whisper ASR transcription process required 46.1 (± 13.9) seconds, while the GPT-4o report generation was the fastest step, completing in an average of 4.9 (± 1.9) seconds. The radiologist editing phase, which involved reviewing and modifying the AI-generated report, took 43.1 (± 34.4) seconds on average.

## Discussion

The findings of this study demonstrate the feasibility of AI-driven transcription and report generation from vernacular-language radiologist-patient conversations for mammography. The AI-generated reports exhibited a high overall concordance rate of 0.94 with radiologist-edited reports, with structured fields such as “Recommendation” and “Comparison” achieving the highest concordance at 0.96. However, discrepancies were noted in more descriptive sections, particularly the “Left Breast Parenchyma” field, which had the lowest concordance rate of 0.64. Error analysis revealed that 50% of the AI-generated reports contained one or more discrepancies, with missed information (16 instances) and incorrect information (13 instances) being the most prevalent errors. Notably, errors were more frequently observed in malignant cases (70%). The majority of these errors were classified as minor or moderate in severity. These results suggest that while this AI-assisted reporting workflow is a potential approach to patient-centered radiology, there is a need for improvement in handling descriptive, free-text sections to enhance overall report quality.

ASR-based tools have been previously explored for transcribing physician-patient interactions and structuring medical documentation, showing promise but lacking large-scale validation (15–17). Evaluations of specialized ASR models for clinical conversations have demonstrated improved transcription accuracy and diarization, with domain-specific adaptations outperforming general-purpose models (16,17). However, no commercially available system has yet been widely adopted (15). Beyond clinical encounters, ASR has also been applied to radiology transcription in non-English languages, with a study adapting the Whisper Large-v2 model for French radiological data achieving a WER of 0.171, highlighting its feasibility for structured documentation in multilingual settings (18).

Prior studies have explored the use of LLM-driven text simplification models to convert complex radiology reports into patient-friendly summaries, making medical findings more understandable to non-specialists (19–21). Studies have shown that these models significantly improve patient comprehension and confidence while effectively reducing reading grade levels, with readability influenced by prompt phrasing (19,20). However, while prior studies have focused on ASR for medical documentation and LLMs for text simplification, research on integrating these technologies to generate structured radiology reports directly from spoken radiologist-patient vernacular conversations remains unexplored. This study presents a novel application of AI-driven transcription and report generation in radiology, introducing an innovative workflow that integrates ASR, LLM-based structured reporting, and human validation to bridge communication gaps in diagnostic imaging.

The implementation of such an AI-assisted workflow has the potential to benefit multiple stakeholders in the healthcare ecosystem. For patients, direct discussion with the radiologist provides a more understandable, accessible, and engaging experience, ensuring they receive direct insights into their imaging findings without reliance on intermediaries (22,23). This can reduce anxiety, improve health literacy, and enable better-informed decision-making. For radiologists, the AI-driven approach facilitates structured patient interactions, providing valuable clinical history, while also allowing for greater efficiency in documentation (24,25). Enhanced patient communication may also lead to greater professional satisfaction, as radiologists can play a more direct and meaningful role in patient care. For referring clinicians, this workflow reduces the burden of explaining imaging findings, thereby allowing physicians to focus on treatment and management decisions rather than radiological interpretation (26,27).

Despite its promise, several challenges need to be addressed before widespread adoption of AI-driven radiology transcription and reporting. Privacy and data security concerns remain paramount, as AI-driven speech recognition involves processing sensitive patient conversations. Future work should focus on developing privacy-preserving AI models that can function locally or on secure hospital servers to mitigate data risks. Additionally, clinical validation in real-world settings is necessary to assess the reliability of AI-generated reports across diverse patient populations, as well as their impact on the patient, radiologist and the referring clinician. Expanding the methodology to other imaging modalities, such as CT, MRI, and ultrasound, could further enhance AI’s impact on patient-centric radiology. Moreover, integrating multimodal AI models capable of analyzing both images and speech transcripts could lead to a more comprehensive AI-assisted diagnostic framework.

This study has certain limitations. First, although we employed detailed prompt engineering to optimize AI responses, we did not fine-tune or retrain any models, which may have limited domain-specific adaptability. Second, the dataset was limited to 50 mammograms, which, while diverse, does not capture the full spectrum of mammographic findings encountered in clinical practice. Additionally, only Hindi-language conversations were evaluated, limiting generalizability to other vernacular languages. Future research should explore multi-language models and validate AI performance across larger datasets and diverse linguistic settings.

In conclusion, this study presents the feasibility of a novel AI-driven workflow that integrates ASR and LLMs to generate structured mammography reports from radiologist-patient conversations in vernacular language. While challenges such as privacy, validation, and scalability remain, this approach represents a significant step toward patient-centric and AI-integrated radiology practice.

## Data Availability

All data produced in the present study are available upon reasonable request to the authors

## Notes

### Competing Interest Statement

The authors have declared no competing interest.

### Funding Statement

This study did not receive any funding

### Author Declarations

Institute Ethics Committee of All India Institute of Medical Sciences, New Delhi gave ethical approval for this work.

